# Characteristic and Outcomes of Patients with ST-segment Elevation Myocardial Infarction who have Non-System Reasons for Delay in Treatment: A Report from the AHA GWTG CAD Registry

**DOI:** 10.1101/2024.11.07.24316939

**Authors:** Tetz C. Lee, James G. Jollis, Haoyun Hong, Tian Jiang, Juan Zhao, Timothy D. Henry, Alice K. Jacobs, Abhinav Goyal, Christopher B. Granger, Kathie Thomas, Jacqueline E Tamis-Holland

## Abstract

**Background:** The past two decades have witnessed reductions in time to diagnosis and reperfusion therapy in patients with ST elevation myocardial infarction (STEMI), largely through improvements in STEMI systems of care. While studies have demonstrated important benefits of timely coronary reperfusion in STEMI patients, those with non-system reasons for delay (NSD) are often excluded from these analyses, limiting insights into the overall quality of care for these patients.

**Methods:** We analyzed the NSD in patients with STEMI undergoing primary PCI who were enrolled in the GWTD-CAD registry from January 1st, 2019, to December 31st, 2021. We examined the patient-level characteristics and outcomes for patients with and without reported NSD. We performed multivariable logistic regression models to examine the association between NSD and in-hospital mortality, adjusting for patient demographics, clinical variables, and social factors. We then categorized hospitals into four groups based on proportion of STEMI patients with NSD and examined the hospital-level characteristics across the quartiles. We further grouped hospitals by quality metric achievement of timely coronary reperfusion and examined the rates of NSD and treatment times for each category of achievement of the quality metric.

**Results:** 74,372 patients were included in the study. 17,741 (23.9%) patients were reported to have NSD. Patients with NSD were older, and more likely to be female, of Black race, and have significant comorbidities including higher rates of cardiac arrest, heart failure and cardiogenic shock on presentation. In-hospital mortality rate was significantly higher in patients with NSD (15.4% vs 2.7%), (Adjusted OR 2.78, [95% CI:2.54-3.04]). Although high-achieving hospitals (those meeting the metrics ≥75%) excluded more patients, they consistently maintained shorter treatment times, even when NSD patients were included in the analysis.

**Conclusion:** NSD in STEMI care is prevalent and linked to higher in-hospital mortality. While concerns about selective case exclusion exist, high-achieving hospitals consistently demonstrated excellent time-to-treatment even when patients with NSD are included.

## Background

The past two decades have witnessed considerable advances in the management of patients with ST elevation myocardial infarction (STEMI) ^1–4^. The American Heart Association’s Mission: Lifeline program, created in 2008, provides a framework for managing these patients across the systems of care and encourages communication and coordination between emergency medical services (EMS), hospital emergency departments and the cardiac catheterization laboratories. The Get With The Guidelines–Coronary Artery Disease (GWTG-CAD) registry was developed to allow health care systems to monitor the time to treatment, therapies provided and hospital outcomes of patients with acute myocardial infarction (AMI). For STEMI patients, this data provides a perspective of management across the EMS and hospital systems. Numerous studies have shown that timely coronary reperfusion in STEMI patients reduces in-hospital and long-term mortality ^5–9^. However, patients with non-system reasons for delay (NSD) were excluded from the most recent and comprehensive analysis^9^.

Exclusion criteria were originally designed to preserve the face validity of quality improvement measures by excluding patients whose treatment times were due to uncontrollable delays. In the case of STEMI care, patients with NSD were not included in the measures of door-to-balloon time for the purposes of public or proprietary quality improvement initiatives. An earlier study based on the CAD-GWTG registry from 2009 to 2011 reported a frequency of NSD of 15% and identified five-fold higher mortality (3% versus 15%) for patients with NSD^10^. The report raised concerns that a significant portion of STEMI patients were excluded from quality improvement assessment and urged that future reports provide a more detailed analyses to potentially expand the focus of the quality-of-care initiatives to include those with NSD. Despite this call to action, the number and frequency of exclusions for NSD has increased over time. In a recent analysis, the proportion of patients with STEMI reported as having NSD in the GWTG-CAD registry increased from 19% to 24% between 2018 and 2021^11^. Notably earlier reports have demonstrated great variation in reporting of NSD on a hospital level ranging from from 0% to 68%, with an interquartile range of 6% to 14%^12^. Studies from matching hospitals noted a two-fold variation in exclusion rates depending on the registry to which the hospital is reporting.^13^ This variation raises the possibility that the approach to quality improvement may rely on identifying reasons for exclusion more than identifying opportunities for improvement^12–14^. Using a national registry of patients with STEMI, we sought to determine the proportion of STEMI patients reported as having NSD and examine the outcomes of these patients. We hypothesized that important opportunities for improved quality of care and outcomes can be identified through a more thorough understanding of NSD.

## Methods

### Data collection

We analyzed the data from the GWTG-CAD registry a national quality improvement registry of patients with acute myocardial infarction. The American Heart Association GWTG-CAD Registry is a voluntary quality improvement program in the United States which has been described previously^9^. Participating hospitals upload clinical data of consecutive patients admitted with ST-elevation myocardial infarction (STEMI) or NSTEMI. Program information and data elements collected in the case report form are available at: https://www.heart.org/en/professional/quality-improvement. Because data were used primarily at the local site for quality improvement, each participating hospital received either human research approval to enroll cases without individual patient consent under the common rule, or a waiver of authorization and exemption from subsequent review by their institutional review board (IRB). Advarra, the IRB for the American Heart Association, determined that this study is exempt from IRB oversight. The data collection and coordination for GTWG programs are managed by IQVIA (Parsippany, New Jersey).

### Study population

For the purposes of this analyses, we examined patients enrolled in the GWTG-CAD registry with acute STEMI referred for primary percutaneous coronary intervention (PCI). Exclusion criteria include patients who received fibrinolysis; who underwent PCI more than 12 hours from symptom onset and were not unstable; or had PCI for NSTEMI. The study evaluated consecutive patients enrolled in the registry between January 1st, 2019, and December 31st, 2021. We grouped patients according to the presence or absence of NSD. We further classified patients with NSD into two groups: those whose reasons were due to cardiac arrest, the need for intubation, or placement of an LV support device, and those whose reasons were unrelated to cardiac arrest, intubation, or the need for LV support device placement.

### Outcomes

The outcomes of interest included time to reperfusion for STEMI and in-hospital mortality. Given that the time to reperfusion varied depending on transportation modes, the outcome measure was defined separately for each presentation mode. First medical contact was defined according to the setting where patients first presented. For those presenting by EMS to a PCI– capable hospital, it was defined as the time they were first evaluated by paramedics; for those who walked in to a PCI-capable hospital, it was defined as the hospital arrival time; and for those requiring interhospital transfer for PCI, it was defined as the time they arrived at the referring hospital. Each outcome were calculated using the published measure defined in STEMI Receiving Center Achievement, Quality, and Reporting Measures (Version 12/2019) ^15^ ^16^. Briefly, the primary objective of the quality program was to provide coronary artery reperfusion (device time) for 75% of patients within 90 minutes of first medical contact for patients initially presenting to PCI-capable hospitals, and 120 minutes for patients requiring hospital transfer for PCI. The goal for each of these outcomes was derived based on the ACC-AHA guidelines.

### Statistical Analyses

To evaluate difference between the patient-level characteristics for patients with and without NSD, we used the chi-square test if appropriate for categorical variables and Kruskal-Wallis test for continuous variables. Multivariable logistic regression modeling using generalized estimating equations was conducted to evaluate if NSD were independent predictors of in-hospital mortality accounting for within-hospital correlation. Patients without reported NSD served as the reference group. The adjusted models included demographics (age, sex, race, and ethnicity) at admission, medical history, and illness severity descriptors such as cardiac arrest prior to arrival, cardiogenic shock on first medical contact (FMC), and heart failure on FMC. We also conducted another adjusted model with adjusting all the above variables plus the social variables including insurance and Social Vulnerability Index (SVI)^17^. These adjusted covariates were used based on a prior study^11^. We reported the in-hospital mortality rates, the odds ratio (OR) in unadjusted and adjusted models. Statistical significance was assessed at a 2-sided α = .05. Patients at sites with missing hospital characteristics were excluded from models. The missing values for clinical continues variables were imputed using multiple imputation techniques (e.g. multiple imputation by chained equations).

For our second analysis, we categorized hospitals into quartiles based on proportion of STEMI patients excluded from their analysis for treatment times due to NSD. We examined hospital level characteristics across these quartiles. Between-group differences were assessed using the chi-square test or Fisher’s exact test if appropriate for categorical variables and Kruskal-Wallis test for continuous variables. We further divided hospitals into four groups according to the percentage of their STEMI cases meeting the guideline recommended time to treatment goals (meeting the metric in ≥ 99% of cases; meeting the metric in 90 % to 98% of cases; meeting the metric in 75% to 89% of cases; and meeting the metric in <75% of cases), and examined the median and interquartile range (IQR) for time to treatment for each of the modes of presentation. The American Heart Association-American College of Cardiology guidelines for STEMI ^4^ emphasize the importance of rapid access to PCI and provide time to treatment goals based on the mode presentation. For patients transported by EMS, the goal is to achieve reperfusion with primary PCI within 90 minutes of first medical contact (FMC). For patients presenting directly to a PCI-capable hospitals (i.e. walk-ins), the Door-to-Balloon (D2B) time should be within 90 minutes from the time they enter the hospital to the initiation of PCI. In cases where a patient initially arrives at a non-PCI-capable hospital, the guidelines recommend transfer to a PCI-capable facility and performing PCI within 120 minutes from the FMC.

Based on the recommendations for time to treatment we looked at three different groups according to their mode of presentation. For each mode of presentation, we first looked at median time to reperfusion for patients who did not have NSD (the standard reporting for GWTG performance metrics). Next, we looked at median time to reperfusion for patients with NSD. Finally, we looked at median time to reperfusion for patients with NSD who did not also have cardiogenic shock, cardiac arrest or the need for an LV support device. All statistical analyses were performed by the AHA Data Science team using R (v4.2.0).

## Results

Between January 1st, 2019 and December 31st, 2021, a. total of 485,339 patients were enrolled in GWTG-CAD registry. By applying the inclusion/exclusion criteria shown in Figure 1, 74,372 patients were finally included in the analysis. Among these 74, 372 patients, 17,741(23.9%) had NSD. The reasons for NSD are depicted in the Supplemental Table 1. Hospitals were able to choose multiple reasons for delay and those reasons shown in the table were not mutually exclusive. The most commonly reported reason for NSD was cardiac arrest and/ or need for intubation, followed by “other reasons” and then difficulty crossing the culprit lesion. The characteristics of patients are shown in Table 1. Patients with NSD were older, and more likely to be female, of black race, and have significant comorbidities. They also had higher rate of cardiac arrest prior to arrival, heart failure on FMC, and cardiogenic shock on FMC; and had a higher social vulnerability index score (were more vulnerable from a socioeconomic standpoint).

**Figure 1.**
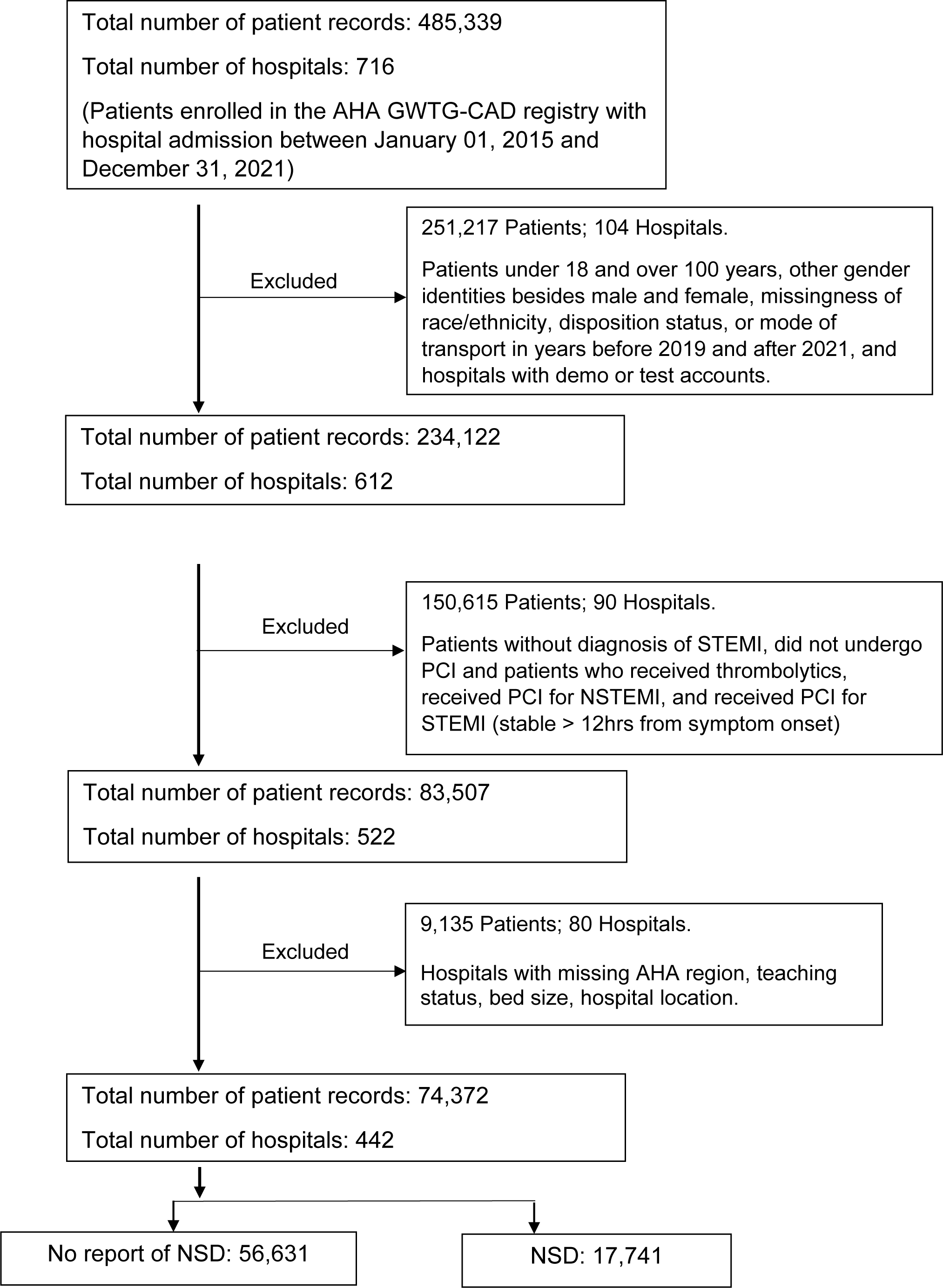
Flowchart of Cohort Selection for Patients with ST-segment Elevation Myocardial Infarction GWTG-CAD, Get With the Guidelines–Coronary Artery Disease registry; STEMI, ST-segment elevation myocardial infarction; NSTEMI, non ST-segment elevation myocardial infarction, PCI, percutaneous coronary intervention; AHA, American Heart Association, NSD, non-system reasons for delay

**Table 1.** Baseline Characteristics of Patients with and without Non-system reason for Delays.

| Variable | No report of Non system Delay | Non-system Delay | Total Patients | P-value |
| --- | --- | --- | --- | --- |
| N (%) | 56631 (76.1) | 17741 (23.9) | 74372 |  |
| Age, years (median, IQR) | 62 (17) | 64 (18) | 62 (17) | <0.001* |
| Female (n, %) | 15420 (27.2) | 5430 (30.6) | 20850 (28.0) | <0.001* |
| **Race/Ethnicity (n, %)** |  |  |  |  |
| Non-Hispanic White | 41838 (73.9) | 12796 (72.1) | 54634 (73.5) | <0.001* |
| Non-Hispanic Black | 5971 (10.5) | 2150 (12.1) | 8121 (10.9) | <0.001* |
| Hispanic | 4459 (7.9) | 1359 (7.7) | 5818 (7.8) | 0.3637 |
| Asian | 2169 (3.8) | 694 (3.9) | 2863 (3.8) | 0.6371 |
| Other | 2194 (3.9) | 742 (4.2) | 2936 (3.9) | 0.0691 |
| **Insurance (n, %)** |  |  |  |  |
| Private/VA/CHAMPUS/Other | 27711 (48.9) | 7777 (43.8) | 35488 (47.7) | <0.001* |
| Medicaid | 5891 (10.4) | 2210 (12.5) | 8101 (10.9) | <0.001* |
| Medicare | 14651 (25.9) | 5305 (29.9) | 19956 (26.8) | <0.001* |
| Self-pay/no insurance | 4602 (8.1) | 1331 (7.5) | 5933 (8.0) | 0.0078* |
| Other/not documented/UTD | 3776 (6.7) | 1118 (6.3) | 4894 (6.6) | 0.0895 |
| **Medical History (n, %)** |  |  |  |  |
| Cerebrovascular disease | 2228 (3.9) | 1076 (6.1) | 3304 (4.4) | <0.001* |
| Diabetes mellitus | 9809 (17.3) | 3606 (20.3) | 13415 (18.0) | <0.001* |
| Currently on dialysis | 288 (0.5) | 202 (1.1) | 490 (0.7) | <0.001* |
| Hypertension | 22798 (40.3) | 8161 (46.0) | 30959 (41.6) | <0.001* |
| Heart failure | 1500 (2.6) | 957 (5.4) | 2457 (3.3) | <0.001* |
| Peripheral artery disease | 1345 (2.4) | 792 (4.5) | 2137 (2.9) | <0.001* |
| Prior Myocardial Infarction | 4806 (8.5) | 1795 (10.1) | 6601 (8.9) | <0.001* |
| Prior Coronary Artery Bypass Graft | 1178 (2.1) | 660 (3.7) | 1838 (2.5) | <0.001* |
| Prior Percutaneous Coronary Intervention | 5931 (10.5) | 2137 (12.0) | 8068 (10.8) | <0.001* |
| Dyslipidemia | 18212 (32.2) | 6133 (34.6) | 24345 (32.7) | <0.001* |
| Active or prior smoking | 21983 (41.1) | 6476 (38.5) | 28459 (40.5) | <0.001* |
| Heart Rate on FMC > 100 (n, %)<sup>†</sup> | 8374 (15.2) | 3619 (21.3) | 11993 (16.6) | <0.001* |
| Systolic Blood Pressure on FMC < 90 (n, %)<sup>†</sup> | 2318 (4.2) | 2501 (14.7) | 4819 (6.7) | <0.001* |
| Initial Serum Creatinine > 1.2 mg/dL (n, %)<sup>†</sup> | 11733 (21.8) | 5747 (34.3) | 17480 (24.7) | <0.001* |
| **Left Ventricular Function Assessment (n, %)<sup>†</sup>** |  |  |  |  |
| < 30 % | 4008 (7.3) | 2590 (15.6) | 6598 (9.3) | <0.001* |
| 30 - 39 % | 8083 (14.8) | 2882 (17.3) | 10965 (14.7) | <0.001* |
| 40 - 49 % | 13471 (24.6) | 3897 (23.4) | 17368 (24.4) | 0.0165* |
| 50 - 59 % | 18793 (34.4) | 4697 (28.3) | 23490 (32.9) | <0.001* |
| 60 - 69 % | 9981 (18.3) | 2432 (14.6) | 12413 (17.4) | <0.001* |
| > 70 % | 330 (0.6) | 126 (0.8) | 456 (0.6) | 0.0332* |
| **Patient Characteristic on FMC (n, %)<sup>†</sup>** |  |  |  |  |
| Cardiac arrest prior to arrival | 1097 (2.4) | 2781 (18.2) | 3878 (6.3) | <0.001* |
| Heart failure on FMC | 3179 (5.8) | 2192 (12.7) | 5371 (7.4) | <0.001* |
| Cardiogenic shock on FMC | 1805 (3.3) | 3238 (18.7) | 5043 (7.0) | <0.001* |
| **Transportation mode to first facility (n, %) -**<br>**Not transferred from another facility** |  |  |  |  |
| EMS -- Air | 796 (1.4) | 252 (1.4) | 1048 (1.4) | 0.9125 |
| EMS -- Ground Ambulance | 27550 (48.6) | 10347 (58.3) | 37897 (51.0) | <0.001 |
| Walk-in | 16936 (29.9) | 3901 (22.0) | 20837 (28.0) | <0.001 |
| **Transferred from another facility (n, %)** | 11349 (20.0) | 3241 (18.3) | 14590 (19.6) | <0.001 |
| Social Vulnerability Index (SVI) percentile rank ≥ 90th (n, %)<sup>†</sup> | 2998 (5.7) | 1140 (6.9) | 4138 (6.0) | <0.001 |

Abbreviations: IQR, interquartile range; FMC, first medical contact; EMS, emergency medical systems.
\*Indicates significant difference with the reference quartile (1st Quartile) by two-sample proportion test, 0.05 as the significance level.
<sup>†</sup> Percentage for variable with missing values are out of total non-missing value counts

In-hospital mortality was significantly higher in those patients with NSD compared to patients without NSD (15.4% vs 2.7%, p< 0.01). Table 2 depicts the unadjusted and adjusted odds for in-hospital mortality among all patients with NSD and for those with NSD unrelated to cardiac arrest, intubation, or the need for LV support device placement. After adjusting for clinical demographics, baseline clinical variables and illness severity descriptors on presentation and SVI, the adjusted odds ratio for in-hospital mortality remain significantly higher for patients with any NSD (adjusted OR, 2.78 [95% CI, 2.54-3.04]) as well as for those with NSD unrelated to cardiac arrest, the need for intubation, or the need for LV support device placement. (Adjusted OR, 1.65 [95% CI, 1.48-1.84]).

**Table 2.** In-hospital Mortality and Risk Adjusted In-Hospital Mortality of Patients with or without non-system reason for delays.

|  | In-hospital mortality (n, %) | Unadjusted |  | Adjusted for age, gender, race/ethnicities, medical history†, cardiac arrest prior to arrival, heart-failure on FMC, cardiogenic shock on FMC, heart rate on FMC, SBP on FMC, creatinine, and LVF assessment |  | Adjusted for age, gender, race/ethnicities, medical history†, cardiac arrest prior to arrival, heart-failure on FMC, cardiogenic shock on FMC, heart rate on FMC, SBP on FMC, creatinine, LVF assessment, Insurance status, and SES (Zip Code: Social vulnerability Index) |  |
| --- | --- | --- | --- | --- | --- | --- | --- |
|  |  | Odds Ratio [95% CI] | P-value | Odds Ratio [95% CI] | P-value | Odds Ratio [95% CI] | P-value |
| No report of non-system reason for delay | 1544 (2.7) | Ref | Ref | Ref | Ref | Ref | Ref |
| Any Non-system reason for delay | 2730 (15.4) | 6.49 [6.00 7.02] | <0.0001 | 2.78 [2.54 3.04] | <0.0001 | 2.78 [2.54 3.04] | <0.0001 |
| NSD unrelated to cardiac arrest, intubation, or the need for LV support device placement† | 731 (6.6) | 2.51 [2.26 2.78] | <0.0001 | 1.66 [1.49 1.85] | <0.0001 | 1.65 [1.48 1.84] | <0.0001 |

Abbreviations: FMC, first medical contact; SBP, systolic blood pressure; LVF, left ventricular function; SES, socioeconomic status, CI, confidence interval.
\* Subjective non-system reason for delay includes difficult vascular access, patient delays in providing consent, difficulty crossing the culprit lesion, need for additional PPE/confirmed infectious disease, and other
† Adjusted medical history variables includes all medical history variables from Table 1

Table 3 depicts the hospital characteristics by quartiles according to the percentage of patients with reported NSD in the said hospital, with Q1 including those hospitals with the lowest proportion of their STEMI patients with NSD and Q4 including hospitals with the highest proportion of patients with NSD. Hospital-level analyses showed wide variations in reporting of patients with NSD from 0% to 100% with an IQR of 17% to 28%. In Q4, the percent of patients with NSD ranged from 28% to 100% of all patients included in the registry with an IQR of 31% to 43%, while the ranges were much narrower in Q1, Q2 and Q3.

**Table 3.** Hospital characteristics grouped by quartiles based on non-systems delays rates for primary percutaneous coronary intervention (4 tiles)

| Variable | All Hospitals | 1st Quartile | 2nd Quartile | 3rd Quartile | 4th Quartile |
| --- | --- | --- | --- | --- | --- |
| N | 442 | 111 | 111 | 110 | 110 |
| Exclusion rate due to NSD (%) (median, [IQR]) | 22.1 (11.4) | 12.5 (5.7) | 19.6 (2.5)† | 24.8 (2.6)† | 37.0 (12.5)† |
| Min – Max | 0.0 – 100.0 | 0.0 – 16.7 | 16.8 – 22.1 | 22.1 – 28.1 | 28.1 – 100.0 |
| 25<sup>th</sup> percentile, 75<sup>th</sup> percentile | 16.8, 28.1 | 8.9, 14.6 | 18.4, 20.9 | 23.4, 26.0 | 30.9, 43.4 |
| **Region (n, %)** |  |  |  |  |  |
| Eastern | 134 (30.3) | 32 (28.8) | 27 (24.3) | 42 (38.2) | 33 (30.0) |
| Midwest | 86 (19.5) | 22 (19.8) | 22 (19.8) | 23 (20.9) | 19 (17.3) |
| Southeast | 75 (17.0) | 21 (18.9) | 22 (19.8) | 11 (10.0) | 21 (19.1) |
| Southwest | 67 (15.2) | 17 (15.3) | 19 (17.1) | 17 (15.5) | 14 (12.7) |
| Western | 80 (18.1) | 19 (17.1) | 21 (18.9) | 17 (15.5) | 23 (20.9) |
| **Teaching Status (n, %)** |  |  |  |  |  |
| Major Teaching Hospital | 74 (16.7) | 17 (15.3) | 12 (10.8) | 20 (18.2) | 25 (22.7) |
| Minor Teaching Hospital | 288 (65.2) | 77 (69.4) | 74 (66.7) | 72 (65.5) | 65 (59.1) |
| Non-teaching Hospital | 80 (18.1) | 17 (15.3) | 25 (22.5) | 18 (16.4) | 20 (18.2) |
| **Ownership (n, %)** |  |  |  |  |  |
| Government | 48 (10.9) | 12 (10.8) | 14 (12.6) | 10 (9.1) | 12 (10.9) |
| Non-profit | 334 (75.6) | 80 (72.1) | 83 (74.8) | 91 (82.7) | 80 (72.7) |
| Private | 60 (13.6) | 19 (17.1) | 14 (12.6) | 9 (8.2) | 18 (16.4) |
| **Hospital Location (n, %)** |  |  |  |  |  |
| Rural | 31 (7.0) | 10 (9.0) | 8 (7.2) | 7 (6.4) | 6 (5.5) |
| Urban | 411 (93.0) | 101 (91.0) | 103 (92.8) | 103 (93.6) | 104 (94.5) |

| Bed Size of Hospital (n, %) |  |  |  |  |  |
| --- | --- | --- | --- | --- | --- |
| 0 – 299 Beds | 247 (55.9) | 63 (56.8) | 73 (65.8) | 60 (54.5) | 51 (46.4) |
| 300 – 499 Beds | 106 (24.0) | 28 (25.2) | 24 (21.6) | 23 (20.9) | 31 (28.2) |
| 500 + Beds | 89 (20.1) | 20 (18.0) | 14 (12.6) | 27 (24.5) | 28 (25.5) |

NSD, non-system reason for delay
The 1st Quartile includes the 25% of hospitals with the lowest NSD rates, and the 4th Quartile includes the 25% of hospitals with the highest NSD rates.
† Indicates significant difference with the reference quartile (1st Quartile) by two-sample proportion test, 0.05 as the significance level.
Continuous values are calculated by Dunn test with Bonferroni correction.

Table 4 depicts the number and proportion of patients with NSD, and the number of hospitals and proportion of NSD categorized by the FMC-related performance metric for each mode of transportation. More patients were treated in hospitals meeting the metrics ≥75% (high-achieving hospitals) than those meeting metrics < 75% (low-achieving hospital). A significantly greater reporting of patients with NSD was noted in the high-achieving hospitals. Median FMC-to-device time were consistently and significantly shorter in high achieving hospitals regardless of the transportation mode (Table 5). High-performing hospitals demonstrated shorter FMC-to-device times, irrespective of the designation of NSD. A shorter median FMC-to-device time was seen when evaluating a group of patients excluding those who have NSD (standard reporting for GWTG Performance Metric); in the group of patients with NSD and in those with NSD unrelated to cardiac arrest and/or the need for intubation or emergent placement of an LV support device.

**Table 4.** Sample size and exclusion rate for each group in hospitals meeting the guideline recommendations for time to treatment metric.

|  | Hospitals meeting the metric <75% | Meeting the metric 75% and < 90% | Meeting the metric >= 90% and < 99% | Meeting the metric >= 99 % |
| --- | --- | --- | --- | --- |
| **EMS patients** |  |  |  |  |
| **PATIENT LEVEL** |  |  |  |  |
| N of Patients | 20382 | 39745 | 11765 | 2418 |
| N of patients with NSD (N, %) | 4187 (20.5) | 9625 (24.2)* | 3248 (27.6)* | 658 (27.2)* |
| N of patients with NSD unrelated to cardiac arrest, intubation, or the need for LV support device placement (N, %) | 2483 (12.2) | 6052 (15.2)* | 2148 (18.3)* | 430 (17.8)* |
| **HOSPITAL LEVEL** |  |  |  |  |
| N of Hospitals | 133 | 201 | 65 | 33 |
| Exclusion rate due to NSD (% , median) | 20.5 (10.3) | 22.3 (10.3)* | 24.1 (12.2)* | 25.0 (18.1) |
| Min – Max | 0.0 – 56.4 | 0.0 – 74.6 | 2.4 – 98.1 | 0.0 – 55.9 |
| 25<sup>th</sup> percentile, 75<sup>th</sup> percentile | 14.7, 25.0 | 17.7, 28.0 | 19.5, 31.7 | 16.7, 34.7 |
| **Walk-in patients** |  |  |  |  |
| **PATIENT LEVEL** |  |  |  |  |
| N of Patients | 1436 | 6155 | 11013 | 2229 |
| N of patients with NSD (N, %) | 229 (15.9) | 1140 (18.5)* | 2022 (18.4)* | 507 (22.7)* |
| N of patients with NSD unrelated to cardiac arrest, intubation, or the need for LV support device placement (N, %) | 168 (11.7) | 875 (14.2)* | 1479 (13.4) | 374 (16.8)* |
| **HOSPITAL LEVEL** |  |  |  |  |
| N of Hospitals | 47 | 124 | 190 | 72 |
| Exclusion rate due to NSD (% , median) | 16.7 (16.8) | 17.8 (13.4) | 17.7 (12.8) | 21.1 (14.6) |
| Min – Max | 0.0 – 52.9 | 0.0 – 70.6 | 0.0 – 57.9 | 0.0 – 100.0 |
| 25<sup>th</sup> percentile, 75<sup>th</sup> percentile | 9.8, 26.6 | 11.3, 24.7 | 11.2, 23.9 | 12.4, 27.1 |
| **Transfer patients** |  |  |  |  |

| **PATIENT LEVEL** |  |  |  |  |
| --- | --- | --- | --- | --- |
| N of Patients | 28430 | 18540 | 7511 | 7661 |
| N of patients with NSD (N, %) | 6410 (22.5) | 4441 (24.0)* | 1885 (25.1)* | 2080 (27.2)* |
| N of patients with NSD unrelated to cardiac arrest, intubation, or the need for LV support device placement (N, %) | 3970 (14.0) | 2847 (15.4)* | 1217 (16.2)* | 1284 (16.8)* |
| **HOSPITAL LEVEL** |  |  |  |  |
| N of Hospitals | 157 | 78 | 30 | 60 |
| Exclusion rate due to NSD (% , median) | 21.8 (11.9) | 21.8 (10.9) | 22.8 (10.4) | 22.6 (14.7) |
| Min – Max | 0.0 – 98.1 | 2.4 – 52.2 | 6.8 – 70.6 | 0.0 – 74.6 |
| 25<sup>th</sup> percentile, 75<sup>th</sup> percentile | 16.1, 28.0 | 16.2, 27.2 | 18.4, 28.8 | 18.3, 33.0 |

Abbreviations: IQR, interquartile range; NSD, non-system reason for delay
\*Indicates significant difference with the reference quartile (1st Quartile) by two-sample proportion, 0.05 as the significance level. Continuous values are calculated by Dunn test with Bonferroni correction.

**Table 5.** Perfusion time (Minutes) for different hospital tiers based on proportion of patients meeting the guideline recommendations for time to <u>treatment metric.</u>.

|  | **Hospital Tiers** |  |  |  |
| --- | --- | --- | --- | --- |
|  | **Meeting the metric**<br>**< 75%** | **Meeting the metric**<br>**≥ 75% and < 90%** | **Meeting the metric**<br>**≥ 90% and < 99%** | **Meeting the**<br>**metric ≥ 99%** |
| **EMS Patients** |  |  |  |  |
| **(N)** | 20382 | 39745 | 11765 | 2418 |
| FMC-to-PCI time excluding patients with NSD (min, median, IQR)<br>(Standard Reporting for GWTG Performance Metrics) | 91 (34) | 81 (29)* | 76 (24)* | 77 (25)* |
| FMC-to-PCI time including patients with a NSD unrelated to cardiac arrest, intubation, or the need for LV support device placement† (min, median, IQR) | 94 (38) | 85 (35)* | 80 (32)* | 80 (34)* |
| FMC-to-PCI time including all patients (min, median, IQR) | 97 (41) | 87 (38)* | 83 (35)* | 83 (41)* |
| **Walk-in Patients** |  |  |  |  |
| **(N)** | 1436 | 6155 | 11013 | 2229 |
| Door-to-device time excluding patients with NSD (min, median, IQR) | 86 (39) | 75 (32)* | 70 (26)* | 67 (25)* |
| Door-to-device time including patients with a NSD unrelated to cardiac arrest, intubation, or the need for LV support device placement† (min, median, IQR) | 89 (45) | 78 (37)* | 73 (29)* | 70 (29)* |
| Door-to-device time including all patients (min, median, IQR) | 90 (45) | 78 (38)* | 73 (31)* | 70 (30)* |
| **Transfer Patients** |  |  |  |  |
| **(N)** | 28430 | 18540 | 7511 | 7661 |
| FMC-to-PCI time excluding patients with NSD (min, median, IQR) | 86 (36) | 80 (31)* | 79 (25)* | 79 (27)* |
| FMC-to-PCI time including patients with a NSD unrelated to cardiac arrest, intubation, or the need for LV support device placement† (min, median, IQR) | 90 (40) | 84 (36)* | 83 (33)* | 82 (31)* |
| FMC-to-PCI time including all patients (min, median, IQR) | 93 (44) | 86 (40)* | 86 (36)* | 84 (34)* |

Abbreviations: IQR, interquartile range; FMC, first medical contact; NSD, non-system reason for delay.
\*Indicates significant difference with the reference quartile (1st Quartile) by two-sample proportion test, 0.05 as the significance level.
†NSD unrelated to cardiac arrest, intubation, or the need for LV support device placement includes difficult vascular access, patient delays in providing consent, difficulty crossing the culprit lesion, need for additional PPE/confirmed infectious disease, and other.

## Discussion

The current study is the most extensive analysis of NSD in patients with STEMI to date. We observed that a large proportion of patients with STEMI have NSD, which is reported in approximately one quarter of patients enrolled in the registry. The most common reported reason for NSD was cardiac arrest and/or the need for intubation. As might be expected, patients with NSD were older and had higher rates of associated comorbid conditions including cardiac arrest, heart failure, and cardiogenic shock on FMC. Patients with NSD had a nearly three times greater in-hospital mortality rate compared to those without NSD, even after adjusting for other co-variables. Notably, delays unrelated to cardiac arrest, the need for intubation, or the need for emergent placement of an LV support device were also associated with a higher adjusted mortality compared with patients without NSD.

Our findings were consistent with previous studies that showed a worse outcome in patients with NSD^10^, and a higher mortality regardless of the reason for delay. Since the early days of PCI, an increase in door-to-balloon time has been correlated with worse outcomes ^7^ ^18^ and this study support these findings. Although these earlier reports are derived from all STEMI population including patients with NSD, quality initiatives that reward hospitals for timely reperfusion often exclude patients with clinical or social factors that could delay treatment, as these factors are viewed as outside the hospital’s capacity to affect change. Yet these patients with NSD (particularly those with cardiogenic shock) are often the ones that derive the greatest benefit from timely reperfusion with data suggesting a correlation between door-to-balloon time and mortality in patients with cardiogenic shock^19^. This information emphasizes the importance of improving treatment times in all patients regardless of the presence of NSD, and the need to shift our focus towards quality improvement efforts that are inclusive of all patients.

Although NSD is more frequently reported in high-achieving hospitals, those high-performing hospitals have also demonstrated shorter median FMC-to-device times across all modes of patient presentation, regardless of the NSD presence. Our findings are novel in that they show high-performing hospitals consistently have better FMC-to-device times among all STEMI population including patients with NSD. This is likely attributable to the vigorous implementation of quality improvement activities, which have been shown to significantly reduce perfusion times and sustain long-term improvements ^20,21^. The fact that these high-achieving hospitals are recognized in the GWTG-CAG program indicates that they consistently execute thorough quality improvement practices. These findings highlight the critical role of ongoing quality improvement efforts in consistently achieving better outcomes for all patient populations.

Our analysis is consistent with an earlier report from the Centers for Medicare and Medicaid Services quality measurement initiative^13^ demonstrating that 28% of patients with STEMI were excluded from the estimation of door-to-balloon time, although the rates of exclusion from the NCDR registry in these same hospitals were much lower (12%) than the rates we report. The rates of NSD we report in the current analysis are also considerably higher when compared to a study from a nationwide registry of STEMI patients hospitalized in 2009 to 2011, which showed that 15% of STEMI patients had NSD^10^. Notably, these rates continue to climb over time. Most recently, the proportion of patients with STEMI excluded from analysis of treatment times due to NSD in the GWTG-CAD registry increased from 19% to 24% between 2018 and 2021^11^. There are a number of reasons to explain this increase in NSD. It is possible that this increase in reported NSD reflects a growing burden of higher risk patients encountered by healthcare system over the years with a larger percentage of patients with vascular disease, complex anatomy, or cardiac arrest/cardiogenic shock referred for PCI in STEMI. Data from the National Inpatient Sample from 2003-2016 has demonstrated increased age, and a growing rate of peripheral vascular disease, and chronic kidney disease cardiogenic shock and other comorbidities among patients undergoing PCI^22^. Furthermore, according to the US National Inpatient Sample from 2004 to 2018, the number of patients hospitalized for cardiogenic shock has tripled^23^. While it is tempting to attribute the rise in reported NSD to the COVID-19 pandemic, which led to increased treatment times due to staffing shortage, and the need for personal protective equipment (PPE), only 7% of patients in this report were coded with need for PPE as the reason for delay.

It is also possible that some of the increase in reporting of NSD is a result of greater administrative efforts to identify a reason for exclusion in those patients with prolonged times to treatment (“gaming of the system”) since the hospitals’ measures of door-to-balloon time are tied to public reporting and pay-for-performance ^13,24,25^. Similar to the previous regional analysis^12^, our hospital-level data showed great variation in exclusion rates (0% to 100%) that appear unlikely to be due to chance or differences in patient characteristics. Notably, a large proportion of patients with NSD had a reason other than the provided explanations. Until recently, the other reasons for delay were not routinely recorded and therefore, we do not have a detailed description of all of the “other” reasons for delay reported by hospitals. However, a small sampling of the recorded reasons for delays based on data collected in more recent years suggest a wide variation in the “definition” used by hospitals to document NSD (Supplemental Table 2). Alternatively, it is possible that high achieving hospitals having the resources and expertise to provide timely care, are also those that manage the sickest patients which accounts for the greater proportion of patients with NSD but also explains the overall shorter reperfusion times, even when patients with NSD are included in the metric. This once again emphasizes the need to evaluate treatment times in all patients, as this may be a better reflection of quality of care provided by each hospital.

This study has several limitations. First, the GWTG-CAD registry is a self-reported registry because the voluntary registry is designed to help participating hospitals to improve their processes of care. Many of the data fields were not required to be completed as a condition of submission, resulting in missing data. We include only patients with full data available, rather than impute missing data fields, as the ability of available fields to predict missing data for highly prognostic terms such as cardiac arrest was quite limited. While we adjusted covariates including demographic characteristics, medical history, clinical presentation, insurance status, and the SVI, it is still possible that unmeasured or residual confounding factors could have influenced the results, potentially leading to an overestimation of the true effect, but not to a degree likely to account for the very large observed mortality differences. Notwithstanding these significant limitations, the data we present here represent 1 of 2 available contemporary registries of STEMI care in the United States that provides an overview of recent practice.

## Conclusions

In conclusion, NSD in patients with STEMI are common, are increasing and are associated with higher in-hospital mortality. Although NSD are more frequently reported in high achieving hospitals, those awarded hospitals have shorter median FMC-to-device time in all modes of patient presentation regardless of the designation of NSD. This information supports the importance of shifting our focus on evaluating and improving treatment times in all patients, regardless of the presence of a reason for delay. We propose that future recognition awards be based on data that is inclusive of all patients in measures of treatment times regardless of the presence or absence of NSD.

## Data Availability

The Precision Medicine Platform is a cloud-based system that allows researchers to collaborate and analyze large datasets from any computer in the world using a secure environment and the power of machine learning. Within the research interface, users have access to assorted datasets, including the industry-changing Get With The Guidelines® registry data to accelerate findings into impactful discoveries. Researchers must have an approved research proposal to access Get With The Guidelines® registry data.

https://precision.heart.org/

https://www.heart.org/en/professional/quality-improvement/get-with-the-guidelines

---

**Supplementary Table 1.** Patient characteristics by types of non-system reason for delays.

| **Variable** | **Difficult Vascular Access** | **Cardiac Arrest and/ or need for intubation** | **Patient delays in providing consent** | **Difficulty crossing the culprit lesion** | **Emergent placement of LV support device** | **Need for additional PPE/confirmed infectious disease** | **Other** |
| --- | --- | --- | --- | --- | --- | --- | --- |
| N | 2203 | 6074 | 743 | 3497 | 943 | 1368 | 5156 |
| Female (n, %) | 833 (37.8) | 1654 (27.2) | 325 (43.7) | 985 (28.2) | 262 (27.8) | 388 (28.4) | 1645 (31.9) |
| Male (n, %) | 1370 (62.2) | 4420 (72.8) | 418 (56.3) | 2512 (71.8) | 681 (72.2) | 980 (71.6) | 3511 (68.1) |
| **Race/Ethnicity (n, %)** |  |  |  |  |  |  |  |
| Non-Hispanic White | 1617 (73.4) | 4422 (72.8) | 508 (68.4) | 2558 (73.1) | 666 (70.6) | 956 (69.9) | 3629 (70.4) |
| Non-Hispanic Black | 261 (11.8) | 719 (11.8) | 96 (12.9) | 473 (13.5) | 102 (10.8) | 169 (12.4) | 595 (11.5) |
| Hispanic | 170 (7.7) | 449 (7.4) | 65 (8.7) | 237 (6.8) | 80 (8.5) | 125 (9.1) | 433 (8.4) |
| Asian | 72 (3.3) | 249 (4.1) | 56 (7.5) | 102 (2.9) | 53 (5.6) | 66 (4.8) | 235 (4.6) |
| Other | 83 (3.8) | 235 (3.9) | 18 (2.4) | 127 (3.6) | 42 (4.5) | 52 (3.8) | 264 (5.1) |

**Supplementary Table 2.** Supplementary delay reason documentation, captured in the 2023 dataset.

| Supplementary delay reason documentation | N |
| --- | --- |
| CT to rule out pulmonary embolism or aortic dissection | 1 |
| Extended transport time | 1 |
| Head CT first because patient hit head on cement | 1 |
| Intra-Aortic Balloon Pump | 1 |
| Initial call was a BLS call, Code 2 | 1 |
| Late presenting STEMI | 1 |
| Traffic | 1 |
| Vessels not expanding and closing | 1 |
| Awaiting family arrival | 1 |
| Phone consultation with cardiovascular surgeon | 1 |
| Septic shock | 1 |

The variable specifying the other reasons is only available in the latest dataset (Jan 1, 2023 to Dec 31, 2023 with cohort size of N=76,191). The majority of non-system delay entries lacked specific reason documentation. This table encompasses all documented delay reasons in our database.

